# SIR-based model with multiple imperfect vaccines

**DOI:** 10.1101/2021.05.07.21256860

**Authors:** Fernando Javier Aguilar-Canto, Ugo Avila Ponce de León, Eric Avila-Vales

**Affiliations:** Facultad de Matemáticas, Universidad Autónoma de Yucatán, Anillo Periférico Norte, Trabaje Catastral 13615, Mérida, C.P. 97119, Yucatán, México; Programa de Doctorado en Ciencias Biológicas, Universidad Nacional Autónoma de México, Mexico City, Mexico

## Abstract

Since the introduction of vaccination in the current COVID-19 outbreak, many countries have approved and implemented vaccination campaigns to mitigate and ultimately curtail the pandemic. Several types of vaccines have been proposed and many of them have finally been approved and used in different countries. The different types of vaccines have different vaccine parameters, and therefore, this situation induces the necessity of modeling mathematically the scenario of multiple imperfect vaccines. In this paper, we introduce a SIR-based model considering different vaccines, and study the basic properties of the model, including the stability of the Disease-Free Equilibrium (DFE), which is locally asymptotically stable if the reproduction number is less than 1. A sequence of further results aims to enumerate the conditions where the reproduction number can be decreased (or increased). Two important mathematical propositions indicate that in general vaccination might not be enough to contain an outbreak and that the addition of new vaccines could be counterproductive if the leakiness parameter is greater than a threshold *η*. This model, despite its simplicity, was validated with data of the COVID-19 pandemic in five countries: Israel, Chile, Germany, Lithuania, and Czech Republic, observing that improvements for the vaccine campaigns can be suggested by the developed theory.

## 1 Introduction

During the ongoing COVID-19 pandemic, caused by the *β*-Coronavirus called Severe Acute Respiratory Syndrome Coronavirus 2 (SARS-CoV-2), multiple strategies have been proposed to control the outbreak, including social distancing, lockdown, travel restrictions, remote schooling [6], most of them with several consequences not only in economic terms [13] but in mental health and social issues [2, 4]. Unfortunately, those measures have not been enough to contain the diffusion of the virus and its corresponding pandemic, therefore, at least 50 organizations started the development of a vaccine against the SARS-CoV-2 [24].

Several compartmental mathematical models have been proposed to evaluate the effectiveness of vaccines to control infectious diseases. Gumel and its collaborators [14] developed a mathematical model incorporating susceptible populations that have been vaccinated and develop the mathematical properties of that system. Other researchers apply concepts of imperfect vaccines and evaluate the behavior on how they control the spread of COVID-19. In this sense, one of the first models with the vaccine compartment which models the case of COVID-19 was proposed in [16]. In [12], the authors considered antiviral controls with combined vaccination, based on an SEIR-like model (SE(Is)(Ih)AR). The relationship between the vaccines and the measures of social distancing and face mask usage is studied in [33]. The majority of the models include only one type of vaccine, but consider different compartments for one and two doses [24, 26, 31].

Most of the literature about compartmental models with vaccination in COVID-19 has focused on the optimal distribution of the vaccines through several human groups. A discussion about the criterion of prioritizing vaccines and their ethical concerns is provided in [27]. A general multigroup SVIR model is studied in [38]. For instance, some papers pursued the approach of optimizing which range of age is better to vaccinate in order to reduce deaths of COVID-19 [6, 23, 24]. Different allocations and prioritization of the vaccines have been studied by literature, including considering essential workers [7, 25], the division of low and high risk groups (defined as the presence of comorbidities) [30], and the social contact network [11].

The diversity of vaccines yields different values in terms of efficacy and other parameters related to imperfect vaccines. This scenario motivates the development of a model which considers the presence of multiple vaccines since many countries have implemented more than one type of vaccine. This article aims to present a SIR-based model, as a first step towards the formulation of models with multiple types of imperfect vaccines. As a preliminary construction of the model and for the sake of simplicity, we only considered compartments *S, I*, and *R* in the model, and the compartments related with the vaccinated individuals *V*_*i*_ with a type of label *i*.

Just a few models have introduced more than one type of vaccine, and general cases are analyzed only by papers such as [29], where the model also includes the new SARS-CoV-2 variants. Rather than constructing a particular model for the case of COVID-19, we would like to emphasize the features of a more general situation and study the possible consequences of a panorama where multiple vaccines are presented and the development of the epidemic follows the structure of the SIR model hypothesis. Hence, a compartment *E* (namely, “exposed”) was not considered and the influence of the new variants might affect the long-term forecasting, nevertheless, more research on the influence of the new variants is needed to quantify the vaccine parameters for these cases.

Despite the simplicity, there are many questions that should be addressed in a mathematical perspective of compartmental models and in the general scenario of multiple imperfect vaccines, before we applied our model in the case of COVID-19 (*a priori*). For example, can an outbreak be stopped only by using vaccines, in all cases but with unlimited resources? Are multiple vaccines better than one type? What strategies might be suggested to reduce the spread of disease when multiple vaccines are available?

After a parameter estimation is completed (*a posteriori*), other questions might arise, including the standard formulations: Are the current vaccine campaign enough to stop the spread of the SARS-CoV-2, at least the first variant? If the vaccine campaign in one country fails to reduce the cases of COVID-19, what strategies can be suggested? What are the effects of the vaccines on the number of infected cases?

This paper consists of three main sections: one devoted to the mathematical model of multiple imperfect vaccines, which is an extension of the so-called VSIR or SVIR model (see [19, 21, 22, 38]); the second section corresponds to the mathematical theoretical results; and finally, we present a parameter estimation for different countries, which are Israel, Chile, Germany, Lithuania, and the Czech Republic. Those countries were only selected because their related data with the model was available online, and was somehow sufficient to perform the fitting of the differential equations. We strongly encourage countries to publish crucial information to allow researchers to study their particular situations. Answers for the first set of questions (*a priori*) are addressed in Section 3 (Mathematical results), particularly in the subsection of *Multiple vaccination theorem*. On the other hand, the second group of questions (related to the empirical results) is studied in Section 4. Finally, we summarized our main results in the *Discussion* and its following *Conclusion*.

## 2 Mathematical models

As stated in the *Introduction*, our model derives from the VSIR model (or SVIR) presented in different papers by authors such as Magpantay and colleagues [19, 21, 22], which are principally focused on childhood vaccination and uses a parameter *p* as the fraction of newborn vaccinated. However, in the case of COVID-19, *p* is set to zero.

Different forms of vaccine failure (and its corresponding parameters) are discussed by the literature [19, 21, 22], including

1. *ε*_*A*_: **Primary vaccine failure**, probability of not getting protected after vaccination.
2. *ε*_*L*_: **“Leakiness”**, probability of getting infected after exposure for a vaccinated individual respecting from an unvaccinated individual.
3. *α*: **Waning rate**, rate of immunity loss across the time.

In [22] the relative infectiousness is considered, but more compartments are required. The waning probability *ε*_*W*_ is another parameter, but is set to 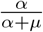. The other parameter related to the vaccination is *r*, the vaccination rate in adults. The set {*ε*_*A*_, *ε*_*L*_, *α, r*} corresponds to the vaccine parameters, the first three depend only on the selected vaccine but *r* is more related to the campaign.

Let *V*_*j*_ the vaccinated individuals with the *j*-th labeled vaccine, *ε*_*j,A*_ ∈ [0, 1) the primary vaccine failure, *ε*_*j,L*_ ∈ [0, 1) the “leakiness”, 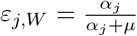 the “waning probability”, *α*_*j*_ ≥ 0 the waning rate of the *j*-th vaccine respectively. The dynamics of *V*_*j*_ are given by

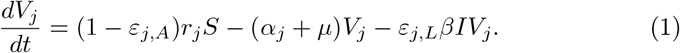

for all *i* = 1, …, *N*, where *β* ≥ 0 the transmission rate and *N* the number of different vaccines. A similar interaction between *V*_*j*_, *S* and *I* is defined in [16], which the vaccination rate as the same parameter and the vaccine efficacy as the leakiness. In practical terms, leakiness will be used as the vaccine efficacy, following the cited paper. The dynamics of the susceptible compartment *S* are given by

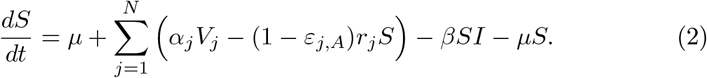

Another possibility of vaccine failure is leakiness, which affects the behavior of *I* (or *E*, if it is considered):

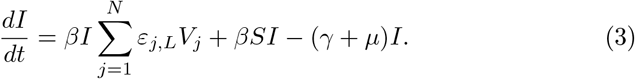

The simplest SIR-based model introduces the dynamics of *R*, given by,

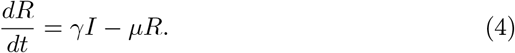

Parameters *β* and *γ* stands for the transmission and recovery rate, whereas *µ* is the birth-death rate for the vital dynamics. A flow diagram of the model is provided in Figure 1.

**Figure 1:**
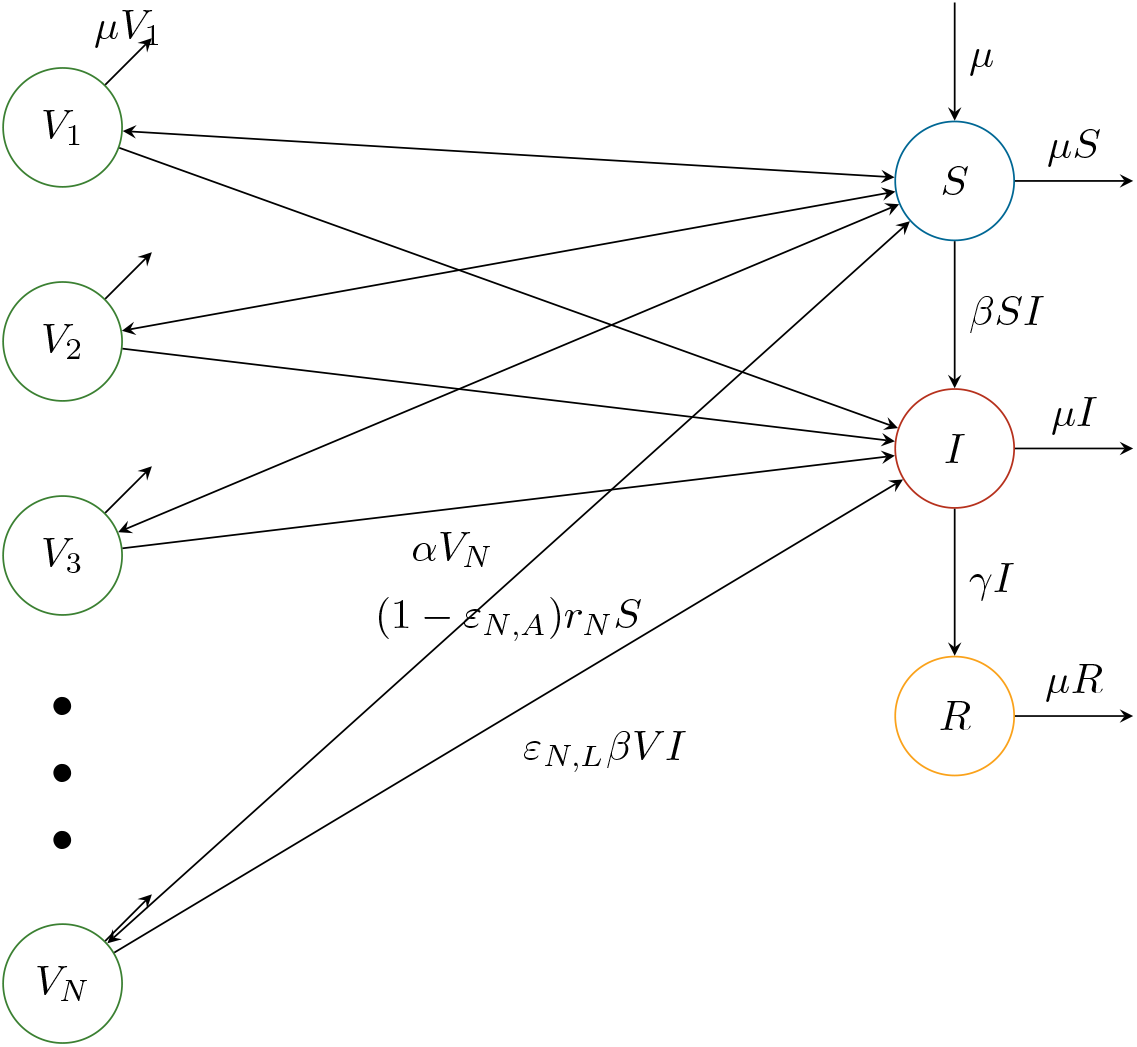
Schematic description of the model (1)-(4).

## 3 Mathematical results

Once the model has been defined, it is needed to verify the basic properties of the model, including aspects related to the stability of the system. In particular, we will focus on the results of the Disease-Free Equilibrium, discuss its stability, and how to reach this value in terms of the variable parameter, which is *r*_*j*_. The other vaccination parameters are considered as fixed, since it is impossible to change them, in terms of public policy, although the addition of new vaccines might be an option. In the following lines, we state and prove the basic properties of the model.

### Proposition 1.

*The model defined by the equations (1)-(4) satisfies the following properties:*

*1.1 The system of equations is invariant in the set* 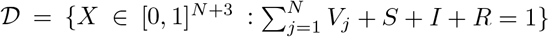.

*1.2 The Disease-Free Equilibrium X*^0^ *of the system is given by*

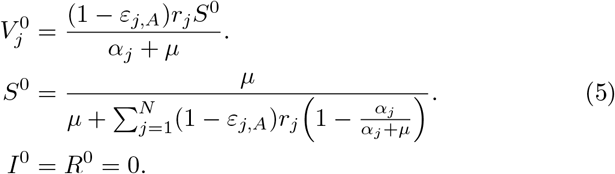

*1.3 The Basic Reproduction Number ℛ*_*c*_ *associated with the model with vaccination is given by*

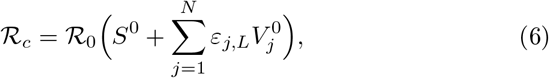

*where* 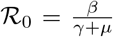 *is the reproduction number of the model without vaccination*.

*Proof*. The proof of 1.1 and 1.2 are standard and will be omitted. In the case of 1.3, using the Next Generation Matrix technique [36], let us consider *x* = (*I, V*_1_, …, *V*_*N*_, *S, R*) and

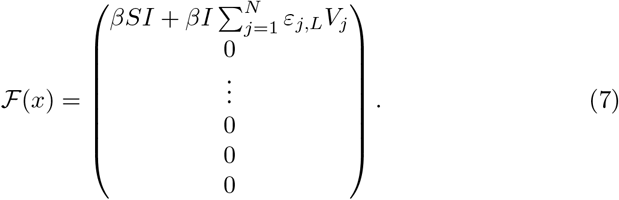

Then, the Jacobian matrix in the DFE *x*^0^ is

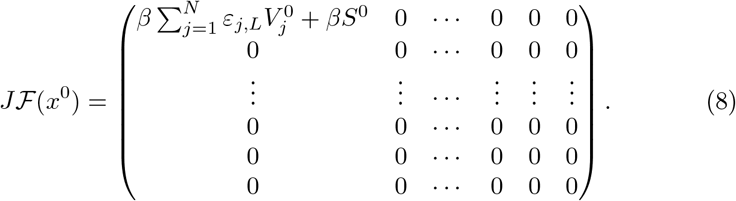

Therefore, 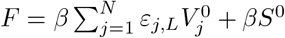 and *V* = *γ* + *µ*, and thus,

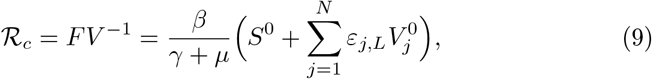

which induces the result, using ℛ_0_, the reproduction number of the SIR model with vital dynamics.

Finally we can discuss the stability of the DFE, which depends on the threshold ℛ_*c*_.

### Proposition 2.

*2*.*1 All Endemic Equilibrium Points x*^*^ *satisfied, for all j* ∈ {1, …, *N*}

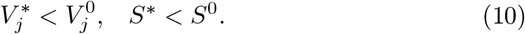

*2*.*2 If ℛ*_*c*_ *<* 1, *the DFE is locally asymptotically stable and the only equilibrium in the system. On the contrary, if ℛ*_*c*_ *>* 1, *the DFE is locally asymptotically unstable*.

*Proof*. In the case of Proposition 2.1, let us suppose that exists *j* such that 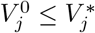 Since

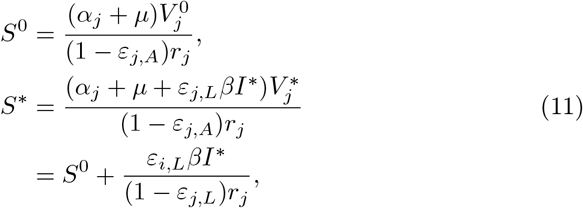

then, *S*^0^ *< S*^*^, which yields 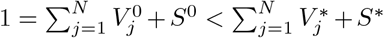, contradicting 2.1. Therefore 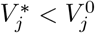,

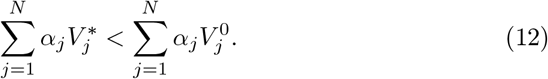

In addition, if *S*^0^ ≤ *S*^*^, since

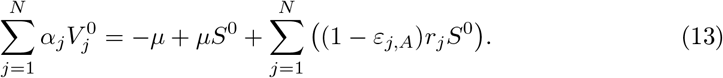

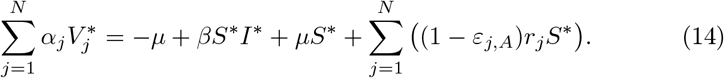

This implies that 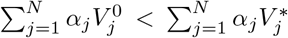, which contradicts (12) and, thus, *S*^*^ *< S*^0^.

Finally, we need to prove 2.2. To proceed, first we require to compute the linearization matrix of the system and evaluate it in the DFE, which is

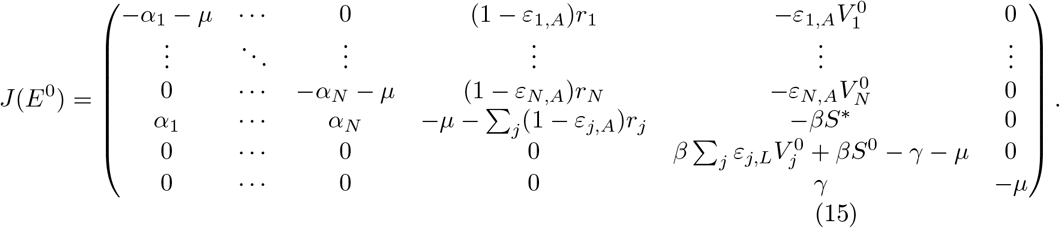

The characteristic equation is given by

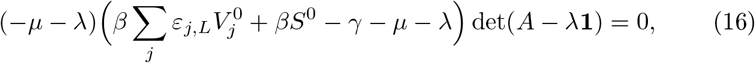

where *A* is given by

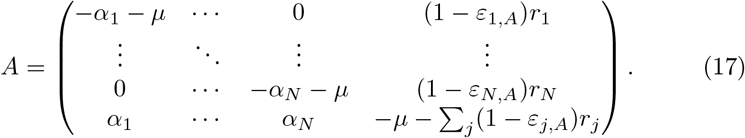

Therefore, *λ*_*N*+3_ = − *µ* and 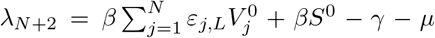. Let *z*_*i*_ = (1 − *ε*_*u,A*_)*r*_*i*_. The other roots are zeros of det(*A* − *λ***1**), which, by Lemma 1 (see *Appendix*), is the polynomial

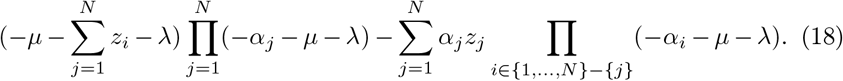

This polynomial has exactly *N* + 1 roots. Let 𝓁_*j*_ = − *α*_*j*_ − *µ*. For the sake of simplicity, let us consider 𝓁_*j*_ sorted in ascending order. Notice that

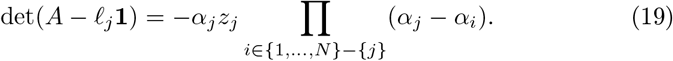

Notice that *α*_*j*_ − *α*_*i*_ *>* 0 if *j < i*. This implies that *j* has *N* − *j* positive products, and, as a consequence, det(*A* − 𝓁_*j*_**1**) has a different sign respecting det(*A* − 𝓁_*j*+1_**1**). By Bolzano’s Theorem, there exists *λ*_*j*_ ∈ (𝓁_*j* − 1_, *𝓁*_*j*_) such that det(*A* − *λ*_*j*_**1**) = 0. Thus, each pair 𝓁_*j*_, *𝓁*_*j*+1_ induces to one solution of det(*A* − *λ***1**) = 0, which give us the zeros *λ*_2_, …, *λ*_*N*_, all of them negative.

Finally, we need to find the remaining roots *λ*_1_ and *λ*_*N*+1_. The leading term of the polynomial is (− 1)^*N*+1^*λ*^*N*+1^. Therefore, lim_*λ*→ − ∞_ det(*A* − *λ***1**) = ∞, but the limit lim_*λ*→ − ∞_ det(*A* − *λ***1**) depends on the parity of *N*. Since for *j* = 1, Π _i ∈ {1,…,N} − {j}_ (*α*_*j*_ − *α*_*i*_) *>* 0, then, det(*A* − 𝓁_1_**1**) *<* 0. Using the Bolzano’s Theorem and the definition of limit, we can can assure the existence of a root *λ*_1_ ∈ (− ∞, *𝓁*_1_).

The remaining root satisfies *λ*_*N*+1_ ∈ ℝ, because if *λ*_*N*+1_ ∈ ℂ − ℝ, then, 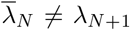 is another complex root. Now, we need to prove that *λ*_*N*+1_ *<* 0. By contradiction, let us suppose that a value *λ* ≥ 0 satisfies det(*A* − *λ***1**) = 0. Thus,

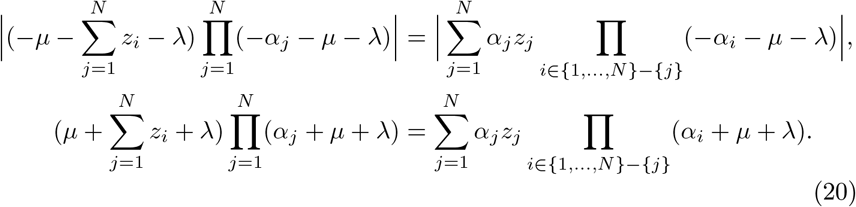

Nevertheless, (20) does not hold, because since *α*_*j*_ + *µ* + *λ > α*_*j*_, therefore

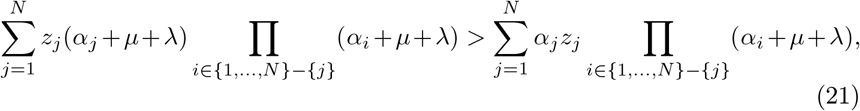

which implies that

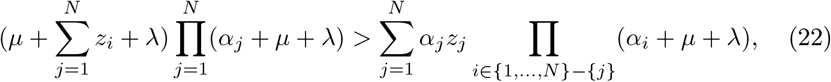

contradicting (20). Now, we have *λ*_1_, …, *λ*_*N*+1_ negative solutions of the original characteristic equation (16). Since the parameters are positive, *λ*_*N*+3_ *<* 0. Notice that, if ℛ_*c*_ *<* 1, then *λ*_*N*+2_ *<* 0, and all the eigenvalues are real and negative, which proves the asymptotical stability of the DFE. On the contrary, if ℛ_*c*_ *>* 1, *λ*_*N*+2_ *>* 0, which implies that the DFE is unstable in this case.

Moreover, if there exists another equilibrium provided ℛ_*c*_ *<* 1, since,

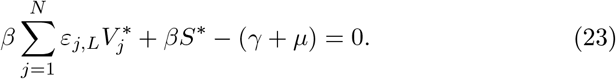

we get 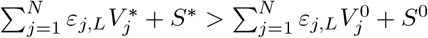, which contradicts 1.1.

### 3.1 Multiple Vaccination Theorem

The following results are related to the basic reproduction number of the model ℛ_*c*_, and the influence of the parameters. Most of them do not require any relevant mathematical tool. However, the importance of the following propositions relies on the interpretative consequences of the model of multiple vaccines. The principal statement in this subsection is given at the end and is called the *Multiple Vaccination Theorem*, which gives some important conditions about the behavior of the function ℛ_*c*_(*r*_1_, …, *r*_*N*_). Complementary results about this function are provided in this section and in the *Appendix*.

#### Proposition 3.

*3.1 Let* 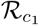 *the Basic Reproduction Number of a model with fixed parameters and* 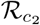 *the Basic Reproduction Number with the same parameters but with* 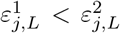 *for some j and where* 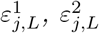 *are the parameters of the first system and second respectly. Then*,

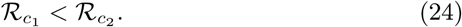

*3.2 Let* 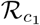 *the Basic Reproduction Number of a model with fixed parameters and* 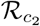 *the Basic Reproduction Number with the same parameters but with* 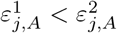 *for some j and where* 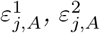 *are the parameters of the first system and second respectly. Then*,

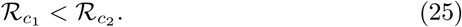

*3.3 Let* 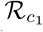 *the Basic Reproduction Number of a model with fixed parameters and* 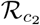 *the Basic Reproduction Number with the same parameters but with* 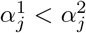 *for some j and where* 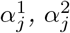 *are the parameters of the first system and second respectively. Then*,

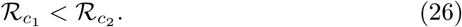

*3.4 Let N* = 1 *and* 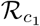 *the Basic Reproduction Number of a model with fixed parameters and* 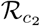 *the Basic Reproduction Number with the same parameters but with r*^1^ *< r*^2^ *for some j and where r*^1^, *r*^2^ *are the parameters of the first system and second respectly. Then*,

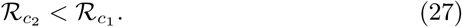

*Sketch*. Proof of 3.1 needs the following remark

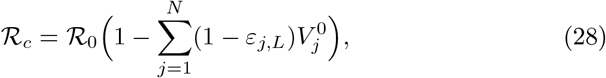

whereas the proof of 3.2 is similar to the next proposition 6 (see *Appendix*). Proposition 3.4 can be proved by computing 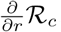, which is,

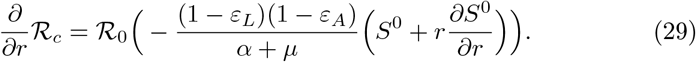

Let *F* =*μ* − (*μp*(1 − *ε* _*A*_)(1 − *ε* _*W*_),

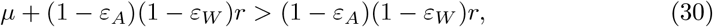

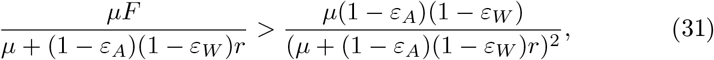

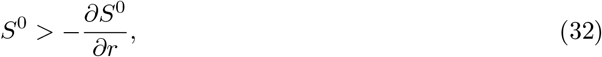

which means that 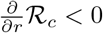. The proof of 3.3 is similar and can be achieved by computing 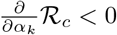

This preliminary result gives us some conditions where the ℛ_*c*_ is decreasing, according to the vaccination parameters. Better vaccine parameters and an increment on the rates yield a minimization over ℛ_*c*_. However, 3.4 is only applicable for *N* = 1, which introduces a fundamental question: What happens for higher values of *N* ? This topic will be discussed at the end of the section. For the following results, we need to introduce the concept of *equivalent vaccines*.

#### Definition 1.

*Two vaccines (labeled with i and j) are said to be equivalent if ε*_*i,L*_ = *ε*_*j,L*_, *ε*_*i,A*_ = *ε*_*j,A*_, *and α*_*i*_ = *α*_*j*_.

Is the vaccination always a secure way to curtail an outbreak? The following propositions gives us a negative answer for this question, which was the first counterintuitive result.

#### Proposition 4.

*The function* ℛ_1_(*r*) *is strictly decreasing and*

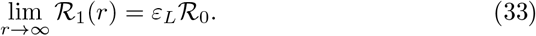

*Proof*. The function is decreasing as shown in Proposition 3.4. In addition, note that lim_*r*→∞_ *S*^0^(*r*) = 0 and since *V* ^0^ + *S*^0^ = 1, lim_*r*→∞_ *V* ^0^ = 1, then, lim_*r*→∞_ ℛ_1_(*r*) = *ε*_*L*_ℛ_0_ holds.

In figure 2 is presented a particular case of the proposition. In this case, is possible to find a value *r* where ℛ_*c*_(*r*) *<* 1. An important corollary of this proposition is the fact that when *ε*_*L*_ *ℛ*_0_ ≥1, no vaccination effort is sufficient to stop the outbreak. A generalization of the last result is given in the following proposition:

**Figure 2:**
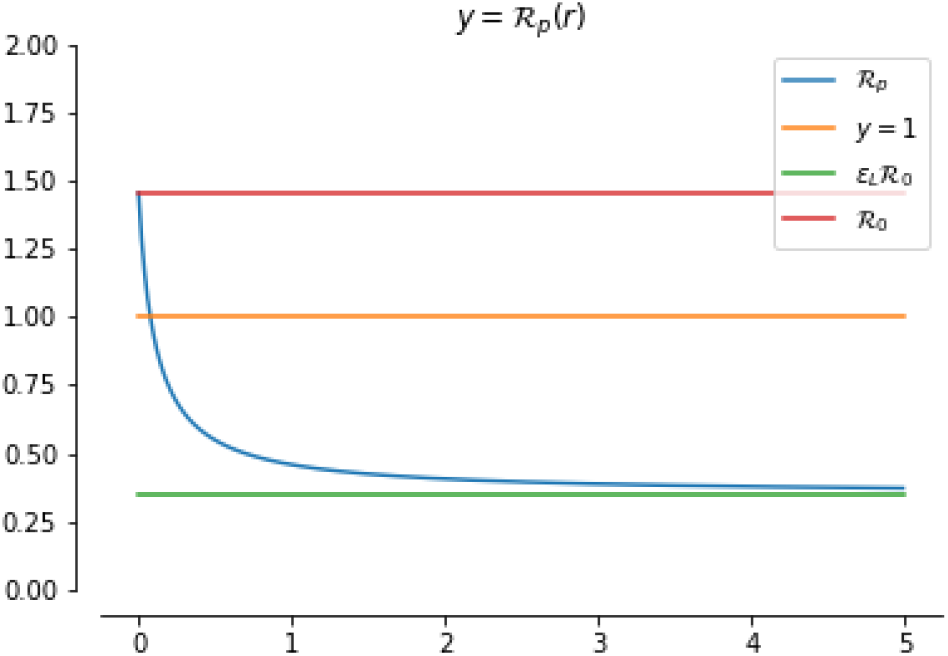
Function ℛ_1_(*r*) with the parameters *ε*_*L*_ = 1 − 0.76, *α* = 0.1, *γ* = 0.1, *β* = 0.16, *ε*_*A*_ = 0.00175, *p* = 0.

#### Proposition 5.

*5*.*1 The function* ℛ_*N*_ (*r*_1_, …, *r*_*N*_) *satisfies*

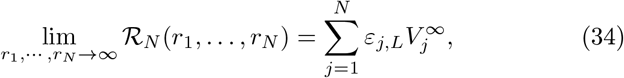

*where* 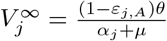 and 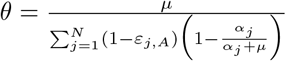.

*5*.*2 For equivalent vaccines*,

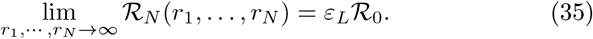

#### Proof.

Preliminary, note that

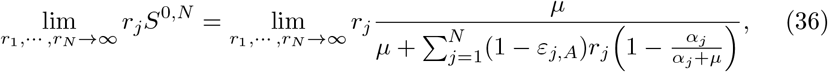

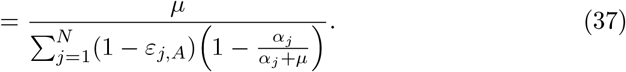

Therefore,

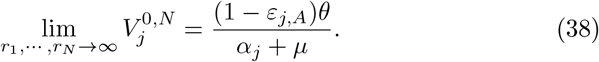

In the case of 5.2,

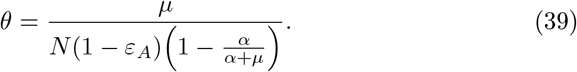

And

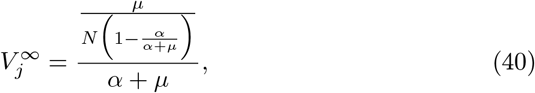

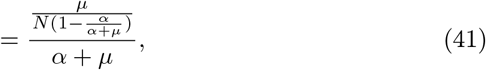

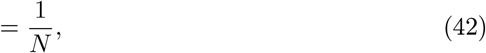

which yields

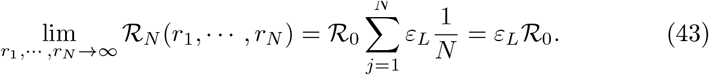

The next mathematical result (Multiple Vaccination Theorem) states that an increment of ℛ_*c*_ can happen if the vaccine parameter *ε*_*N,L*_ of an added vaccine is not low enough. Although is possible to believe that an increment in the vaccination rates is always beneficial in terms of ℛ_*c*_ optimization this might not always happen. In figure 3, we see an example where the increment of *r*_2_ yields an increment in ℛ_*c*_, which is also critical in this case, because ℛ_*c*_(*r*_2_) *>* 1 for some values of *r*_2_. However, if *ε*_2,*L*_ = 0.4, the problem is solved.

**Figure 3:**
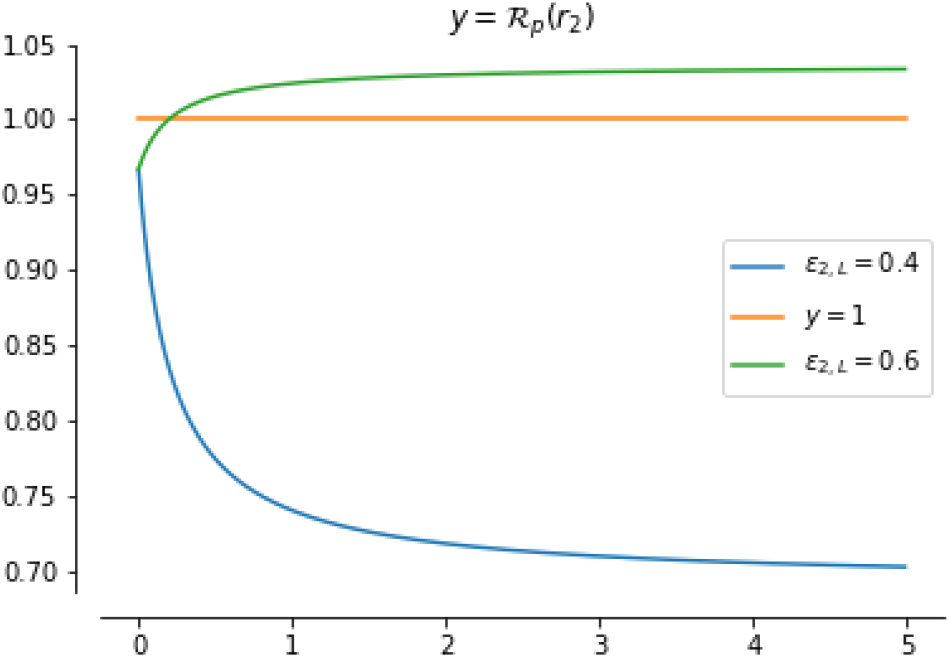
Function ℛ_1_(*r*_2_) with the parameters *ε*_1,*L*_ = 0.05, *α*_1_ = *α*_2_ = 0.1, *γ* = 0.1, *β* = 0.19, and *ε*_1,*A*_ = *ε*_2,*A*_ = 0.05. In the blue line, we set *ε*_2,*L*_ = 0.4 and in the green line, *ε*_2,*L*_ = 0.6. *r*_1_ is fixed to 0.1.

The last reasoning gives us clues about the relationship between *ε*_*j,L*_ and ℛ_*c*_. In Proposition 6 of the Appendix, we can see that if *ε*_*j,L*_ = 0, the reduction is guaranteed. In the following theorem, we state that *ε*_*j,L*_ acts as a unique critical parameter providing the conditions for the desired reduction.

#### Theorem 1

(Multiple Vaccination Theorem). *Let u such that ℛ*_0_*u is the reproduction number of the model with N* – 1 *vaccines, ℛ*_*c*_ *the reproduction number of the model with N vaccines. Let*

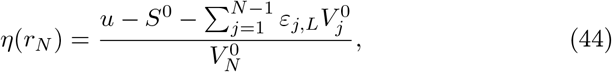

*then*,

1. *η*(*r*_*N*_) = *η*(1) *for any choice of r*_*N*_ *>* 0. *Thus, η is a constant value for r*_*N*_ *variable. η is also constant over any changes of the vaccines parameters of the N -vaccine*.
2. *If ε*_*N,L*_ *< η*, ℛ_*c*_(*r*_*N*_) *is a decreasing function and* ℛ_*c*_(*r*_*N*_) *<* ℛ_0_*u*.
3. *If ε*_*N,L*_ = *η*, ℛ_*c*_(*r*_*N*_) *is constant and* ℛ_*c*_(*r*_*N*_) = ℛ_0_*u*.
4. *If ε*_*N,L*_ *> η*, ℛ_*c*_(*r*_*N*_) *is an increasing function, and* ℛ_*c*_(*r*_*N*_) *>* ℛ_0_*u*.

#### Proof.

Preliminary, note that

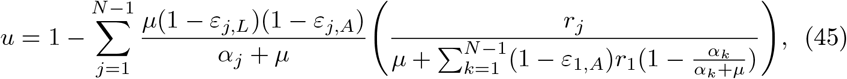

and

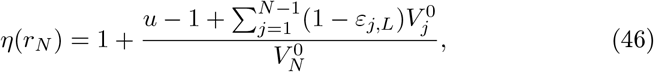

Reducing the expressions, let *B*_*j*_ = (1 − *ε*_*j,A*_)(1 − *ε*_*j,W*_)

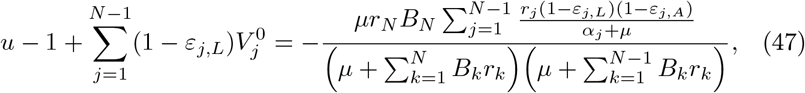

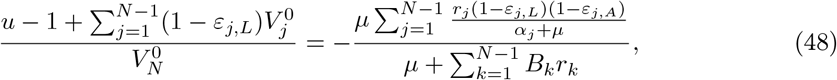

which proves that *η* is constant over any changes of *r*_*N*_, *ε*_*N,A*_ and *α*_*N*_. On the other hand, the line equation

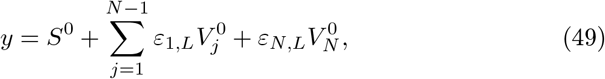

is equal to *u* if *ε*_*N,L*_ = *η*. If 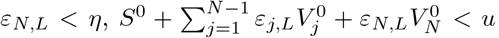, which implies that ℛ_*c*_(*r*_*N*_) *<* ℛ_0_*u*. Similarly, if *ε*_*N,L*_ *> η*, ℛ_*c*_(*r*_*N*_) *>* ℛ_0_*u*.

Finally, differentiating ℛ_*c*_ we get

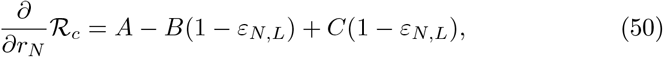

where,

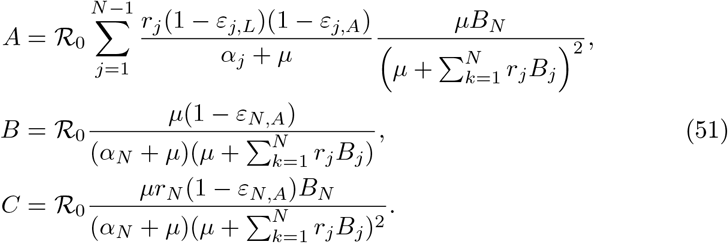

Then,

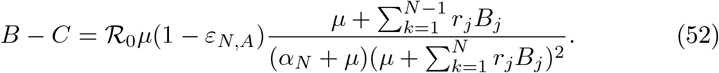

and,

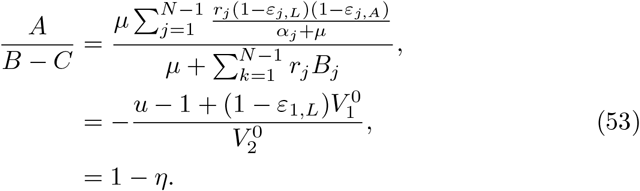

If *ε*_*N,L*_ *< η*, 1 − *ε*_*N,L*_ *>* 1 − *η*,

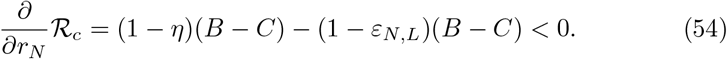

which yields that ℛ_*c*_(*r*_*N*_) is decreasing. The result is analogous for the remaining cases.

Since ℛ_0_*u* is the reproduction number of the system with the first *N* − 1 vaccines, the *N* -th vaccine might be added only if *ε*_*N,L*_ *< η*. Otherwise, ℛ_*c*_ will increase or remain intact. Some remarks must be commented about the Multiple Vaccination Theorem (MVT). It is a criterion about when a new type of vaccine can be added. In Figure 3 we see that *ε*_1,*L*_ *< η < ε*_2,*L*_, but it should be warned that *η* is not a fixed value and instead, it depends on the parameters of the previous vaccines, including the rates. If a country selects vaccines with lower parameters *ε*_*j,L*_, *ε*_*j,A*_,*α*_*j*_ with a insufficient rate, the addition of a new vaccine with higher parameters could produce the opposite effect as the expected.

If ℛ_*c*_ grows after the addition of a new vaccine, this means that the vaccine should not have been added, and rather, an increment on the previously applied vaccines could be a better strategy. It is important to say that the MVT is useful to analyze the behavior of the vaccination rates. Proposition 3.4 indicates that any increment of the rates with *N* = 1 yields a decrement on ℛ_*c*_. As a consequence, the method could study the case with only one vaccine, with the lower leakiness, and then, consider the addition of the next vaccines. If a vaccine has insufficient leakiness, then it should be discarded since it is not helping to reduce the reproductive number. This procedure can be used to determine how to adjust the vaccination rates to minimize ℛ_*c*_(*r*_1_, …, *r*_*N*_), and when to approve a new vaccine.

## 4 Empirical results

The first consideration about parameter estimation of the model is the fact that vaccine parameters are fixed and given by the previous studies (see table 1). Parameters *β, γ* and *µ* are unknown, thus, must be fitted with data. Most vaccine parameters are provided in Table 1 (*ε*_*j,L*_, *ε*_*j,A*_, *w*), but *r*_*j*_ needs to be taken from data. Particularly, *r*_*j*_ must satisfy

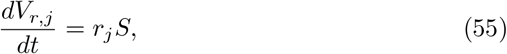

where *V*_*r,j*_ is the compartment of the reported vaccinated. This group is slightly different from *V*_*j*_ since one vaccinated individual might have experience vaccine failure and not be considered in *V*_*j*_. The susceptible *S* and *V*_*r,j*_ are both given in the datasets. However, in the case of multiple vaccines, vaccination is considered as the completion of the required doses, and the datasets provide the vaccinations by manufacturer and the number of the vaccinated people with complete doses. We used the fraction of the vaccinations by the manufacturer (number of applied doses of type A divided by the number of total doses applied) to estimate the actual number of complete vaccines by manufacturer, which was the required data. This estimation could derive some problems, for instance, the estimated cumulative cases of vaccinated individuals by type are not always increasing, although the decrements are small and can be statistically dismissed.

**Table 1:** Parameters of the model. The waning rate remains unknown for all cases but CanSino (6 months). Nevertheless, since estimations are considering a window of fewer than six months, in this paper we considered *α*_*j*_ = 0 for all *j*.

| Vaccine | $1 - \varepsilon_{i,L}$ | $\varepsilon_{i,A}$ |
| --- | --- | --- |
| BNT162B2 (Pfizer) | 0.95 ([0.903, 0.976]) [28] | $3.77 \times 10^{-4}$ [15] |
| Moderna (mRNA-1273) | 0.941 ([0.893, 0.968]) [3] | $7.23 \times 10^{-4}$ [3] |
| ChAdOx1 nCoV-19 vaccine<br>(AZD1222, AstraZeneca) | 0.76 [0.68, 0.82] [37] | 0.00268 [15] |
| Ad26.COV2.S<br>(Johnson & Johnson) | 0.663 ([0.599, 0.718]) [18] | 0.0091 [18] |
| Ad5-nCoV (CanSino) | 0.657 [10] | 0.00175 [15] |
| SinoVac | 0.5065 [8] | 0.0206 [8] |

Following [32], we optimize the parameters *β* and *γ* of the system by using the loss function

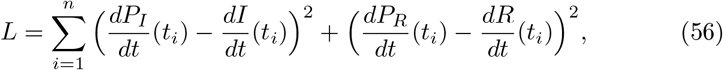

where *P*_*I*_, and *P*_*R*_ are the outputs of the polynomial regression (with a degree equal to 10) with the reported *I* and *R. L* was optimized using gradient descent. Since this method minimizes the error between the derivatives, a final refinement via grid search [1] was performed with the norm ‖*x*‖ _1_ (sum of the absolute difference between prediction and data). For simplicity, *r*_*j*_ and *µ* were also fitted by using grid search.

### 4.1 Data collection

Data was collected from two different sources:

- Records from reported recovered and cumulative infected were taken from the John Hopkins repository [35], widely used in research.
- For vaccination data, we used the repository of *Our World in Data* [17], including the datasets of vaccines per country and vaccines by the manufacturer. The last dataset was restricted for just a few countries, which were the only ones considered in this context.

The first date for optimization purposes was selected according to the presence of all vaccines at a significant rate. All parameter estimations ended on May 1, 2021.

### 4.2 Israel

The first date for the estimations in the case of Israel is January 12, 2021. Pfizer was the only vaccine considered because the usage of Moderna (the other type of vaccine bought by this country) is scarce [34]. Table 2 provides the estimated parameters with the model with one vaccine (VSIR). It was interesting to find that ℛ_0_ *>* 1, but ℛ_*c*_ *<* 1, showing convergence towards the DFE. An confidence interval for ℛ_*c*_ using the intervals for the vaccine parameters is given by (0.05507, 0.14419). As we shall see, this situation is not unique for Israel.

**Table 2:** Estimated parameters for Israel.

| Parameter | Value |
| --- | --- |
| $\beta$ | 0.11808 |
| $\gamma$ | 0.09449 |
| $\mu$ | 0.00023 |
| $r$ | 0.0109 |
| $\mathcal{R}_0$ | 1.24665 |
| $\mathcal{R}_c$ | 0.08682 |

Figure 4 presents the graphs of the behavior of the outbreak in Israel, and its approximation with the model. Perhaps the main problem relies on the fact that the vaccination rate is not constant and a reduction in this rate is evident in the final days. Dynamic *r* might be more convenient for this particular case, for instance, a logistic-like model.

**Figure 4:**
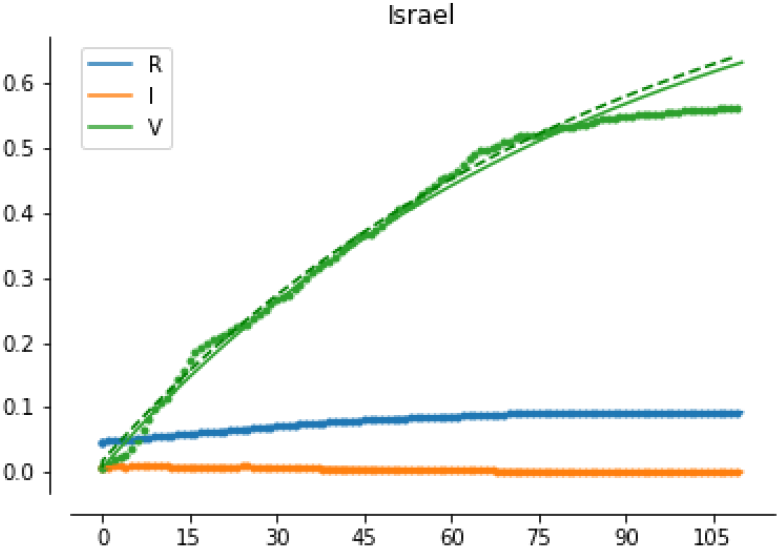
Development of the dynamics of COVID-19 in Israel with vaccination. The dotted green line represents the reported vaccinated.

Critical value *η* = 0.06964, according to the estimated parameters. The theory (specifically Theorem 1) described previously establishes that Israel should not add any vaccine with *ε*_*L*_ *> η*. Surprisingly, this includes most of the vaccines excluding Moderna, which is used in fewer quantities. In this sense, Israel can continue with its successful strategy and should avoid inclusions of other types of vaccine.

### 4.3 Chile

In this case, the initial date was set to February 27, 2021. In this case, two vaccines were implemented: Pfizer and SinoVac. Main results are condensed in Table 3 and Figure 5. Despite suffering a recent peak of new infections, this parameter estimation shows that this country might manage to continue reducing the cases of COVID-19. Again, in this case, ℛ_0_ *>* 1 *> ℛ*_*c*_, which indicates that the vaccination campaign has successfully covered the requirements to contain the outbreak. A confidence interval of ℛ_*c*_ is given by (0.60845, 0.61397).

**Table 3:** Estimated parameters for Chile.

| Parameter | Value |
| --- | --- |
| $\beta$ | 0.19016 |
| $\gamma$ | 0.15159 |
| $\mu$ | 0.0003 |
| $r_1$ | 0.00044 |
| $r_2$ | 0.00667 |
| $\mathcal{R}_0$ | 1.25194 |
| $\mathcal{R}_c$ | 0.61041 |

**Figure 5:**
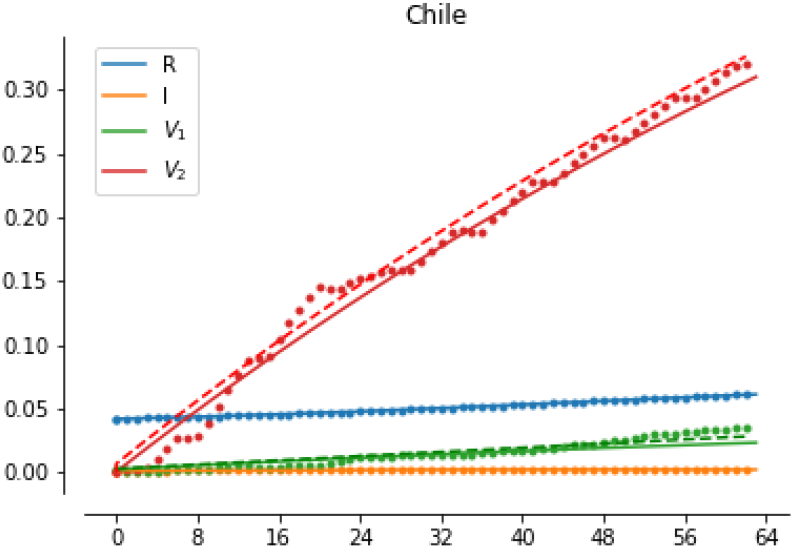
Development of the dynamics of COVID-19 in Chile with vaccination.

Nevertheless, it is important to say that a computation of the threshold *η* with the parameters of the first vaccine (BNT162B2 of Pfizer) yields 0.43522, which is lower than 1 − *ε*_2,*L*_. This shows a case where the application of the second type of vaccine is unnecessary: without this application the value of ℛ_*c*_ would be 0.54488, which is lower that the actual value. Luckily, this does not seem to affect the final situation of Chile.

### 4.4 Germany

Three vaccines were primarily used in Germany: Pfizer, Moderna, and AstraZeneca labeled as 1, 2, 3 respectively. The initial date was February 27, 2021. The results of the parameter estimation are presented in Table 4 and Figure 6. The critical threshold *η* is equal to 0.88337 for the model with only one vaccine (Pfizer), and 0.8715 for the model with two (Pfizer-Moderna). A confidence interval of ℛ_*c*_ using the provided intervals of the leakiness of the vaccines is (0.98682, 1.00833), and the possibility of ℛ_*c*_ ≥1 might not be excluded.

**Table 4:**
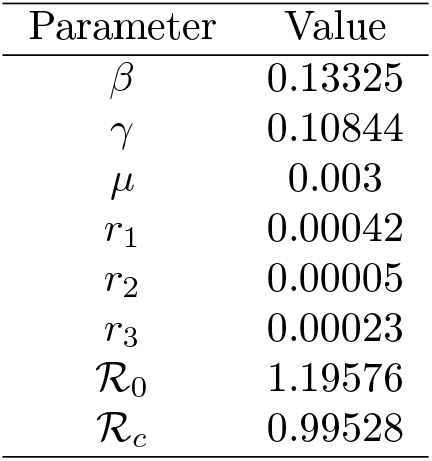
Estimated parameters for Germany.

**Figure 6:**
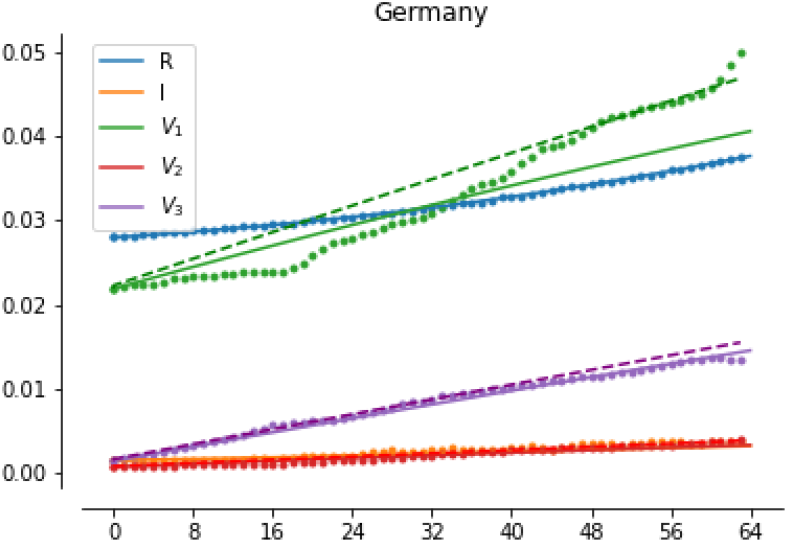
Development of the dynamics of COVID-19 in Germany with vaccination.

In both cases, the remaining vaccines have a leakiness lower than *η*, which confirms that the selection of the strategy does not result in an increment in the reproduction number, which is almost 1. This discards the selected vaccines as a factor on the relatively high ℛ_*c*_, and by ridding one of the types of vaccines, no improvement could be observed if the vaccination rates of the remaining vaccines are kept constant. Moreover, any increment on the rate of the vaccines can help to decrease the reproduction number. In this case, the vaccination rates seem to be slow, thus, public policy should focus on this aspect. Finally, we should note that *η* = 0.83234 for the three vaccines used, which means that any vaccine can be added to minimize the reproduction number.

### 4.5 Lithuania

The three European countries considered in this paper have used the same three vaccines: Pfizer, Moderna, and AstraZeneca, using the same labels. In the case of Lithuania, the first date of analysis is February 9, 2021. The results are summarized in Table 5 and Figure 7. The confidence interval of the reproductive number is (0.72226, 0.77301).

**Table 5:** Estimated parameters for Lithuania.

| Parameter | Value |
| --- | --- |
| $\beta$ | 0.0668 |
| $\gamma$ | 0.05466 |
| $\mu$ | 0.001005 |
| $r_1$ | 0.0004 |
| $r_2$ | 0.00009 |
| $r_3$ | 0.00028 |
| $\mathcal{R}_0$ | 1.1999 |
| $\mathcal{R}_c$ | 0.74219 |

**Figure 7:**
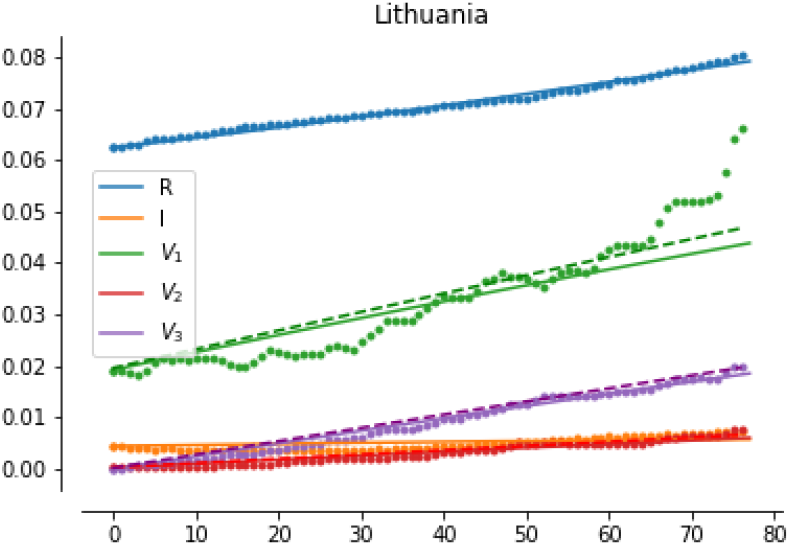
Development of the dynamics of COVID-19 in Lithuania with vaccination.

Just like the case of Germany, two thresholds *η* were computed: using solely the Pfizer vaccine, which was 0.72961, and with Pfizer and Moderna, which yielded 0.68926. Usage of AstraZeneca is, therefore, justified and helps in reducing the reproduction number. Finally *η* = 0.61854 includes the three vaccines, and shows that the addition of vaccines with lower vaccine efficacy is not advised, but, again, all the considered vaccines can be useful in the task of reducing ℛ_*c*_.

### 4.6 Czechia

Four vaccines were implemented in Czechia: Pfizer, Moderna, AstraZeneca, and Johnson & Johnson. Since the vaccination with Johnson & Johnson has started on April 22, 2021, and just a few doses have been applied, we did not consider this type. The selected initial date for analysis is February 11, 2021. Results are provided in in Table 6 and Figure 8 and the confidence interval of ℛ_*c*_ is (0.0617, 0.16309).

**Table 6:** Estimated parameters for Czech Republic.

| Parameter | Value |
| --- | --- |
| $\beta$ | 0.09512 |
| $\gamma$ | 0.07877 |
| $\mu$ | 0.000001 |
| $r_1$ | 0.00071 |
| $r_2$ | 0.00012 |
| $r_3$ | 0.00016 |
| $\mathcal{R}_0$ | 1.20756 |
| $\mathcal{R}_c$ | 0.09983 |

**Figure 8:**
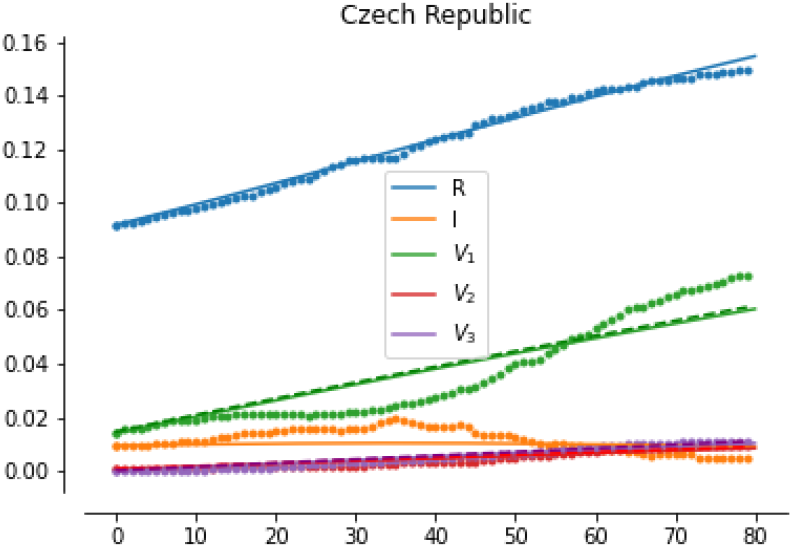
Development of the dynamics of COVID-19 in Czech Republic with vaccination.

Even when the vaccination campaign shows good results, the critical value *η* = 0.05134 for the vaccine Pfizer, which is interesting low, but it implies that the addition of the types Moderna and AstraZeneca was not needed. Value *η* with the model with two vaccines is 0.05244, which confirms that the addition of AstraZeneca was not required, as well as the addition of Moderna.

## 5 Discussion

Vaccination has been described as the best hope against the current pandemic of COVID-19, and possibly, against future important outbreaks [14, 23]. When we use imperfect vaccines (in terms of efficacy) to face a pandemic and more than one type of vaccine is available, many questions arise around the possibility of curtailing the outbreak and the best strategy in the vaccination campaign. Mathematical modeling simplifies the dynamics of the vaccination and the epidemic to simulate, anticipate, predict, and answer the important questions about the topic.

Does this model apply to the current pandemic of COVID-19? In a first advance, the model is motivated by the existence of several types of vaccines, but the inclusion of a SIR model with vital dynamics (which is when *N* = 0) seems to be too simple to describe the current dynamics of COVID-19. There are many other reasons why a more complex model can improve in forecasting future scenarios, but, for instance, the behavior of the rate of vaccinations is not always easy to model, since depends on public policy and sudden changes might be done. Some possible sources of error in the proposed model are listed here:

1. Outbreak parameters *β* and *γ* might vary across the time.
2. Vaccination rates *r*_*j*_ are not constant.
3. Reported recovered individuals might be susceptible again.
4. Most vaccines require two doses (another compartment is required).
5. Relaxation of the measures after vaccination.
6. New SARS-CoV-2 variants might resist vaccination [20].

Different approaches have been proposed to face these difficulties, as presented in the introduction, at least for items 4-6 of the previous list. For long-term forecasting, it is harder to deal with the mentioned problems. For example, the SIRD with dynamical parameters of Caccavo [9] successfully described the outbreak in Italy for a period, but the peak of infection was not April 17th, 2020. Similar cases can be found in the literature, and therefore, it is important to consider that the compartmental model usually presents limitations for long-term prediction (even more than two months). Despite it, parameter estimation with the data is needed to validate this model, even when further considerations will be added to improve the quality of the fittings.

COVID-19 motivates the generalization of the VSIR model with multiple vaccines, but instead of only focusing on this disease, we provided a more general preliminary model, but excluding the compartments of the extended models such as SEIR and SIRD. In this manner, our model attempts to use only the necessary compartments to extend the SIR model with multiple types of vaccines. However, the addition of *E, D* and other compartments, time-dependent parameters, and specific compartments for one and two doses would improve the model for the sole study of the COVID-19 pandemic.

Theoretical analysis showed two important results. Could the vaccination itself manage to eliminate the outbreak? Proposition 4 and its general version 5 provide a negative answer for this question: if the reproduction number and the leakiness of the vaccine are high enough it is impossible to reach ℛ_*c*_ *<* 1, even with a very high vaccination rate. At the beginning of the pandemic, Billah, Mamun Mian, and Nuruzzaman Khan [5] estimated a summary for the reproductive number, which was 2.87. In this manner, vaccines with *ε*_*L*_ higher than 0.34843 might reduce the reproductive number but not enough to curtail the outbreak. However, the reproductive number is different across the countries and time, and nowadays it seems to be lower for most countries.

Another important proposition is provided in Theorem 1 (Multiple Vaccination Theorem), which states that exists a threshold *η* for a model with *N* − 1 vaccines such that, if the leakiness *ε*_*N,L*_ *< η*, any increment on the rate *r*_*N*_ results in a decrement on the reproductive number. This was quite expected, but the unusual part of the theorem says that if *ε*_*N,L*_ = *η*, the increments on the rate do not change the reproductive number, and a worse situation is observed if *ε*_*N,L*_ *> η* when the increments on the rate yield an increment on ℛ_*c*_. Therefore, public measures might consider the main findings and establish criteria to add new types of vaccines or reject previously approved vaccines. It is important to mention that all imperfect vaccines reduce the baseline reproduction number ℛ_0_, but if the addition of one vaccine is increasing the value of ℛ_*c*_, it should be retired.

Parameters of the model were fitted using the available data of five countries: Israel, Chile, Germany, Lithuania, and the Czech Republic. In general, it was possible to perform the parameter estimation, but there are some limitations of the model that we should be aware of. First of all, a SIR-based model might be too simple for the current dynamics of COVID-19, and in general, this model fails to capture the characteristic waves of this particular pandemic. The spread of new variants of SARS-CoV-2 is another source of problems since the vaccines have not been extensively tested for these new strains. Thus, even when a model indicates that ℛ_*c*_ *<* 1 and data show the end of the outbreak, the model does not consider the scenario when protection fails for these new variants. Finally, another main problem was the fact that the vaccination rates are rarely constant.

For all the studied countries, ℛ_*c*_ *<* 1, but in the case of Germany, we can not assure with a confidence of 95 % that ℛ_*c*_ *<* 1. Analysis of the critical value *η* showed that two countries (Chile and Czech Republic), implemented vaccines that are increasing the reproduction number. Moreover, Germany can receive the remaining vaccines from Chile and Czechia, and reduce its reproductive number. This means that a better distribution of the vaccines can be performed if we consider the mathematical results.

## 6 Conclusions and further work

The main topic of this paper consists of an extension of the classical SIR with vital dynamics towards the inclusion of different types of vaccines, which is mainly motivated by the current situation of the pandemic of COVID-19. Theoretical analysis is performed on the proposed model, principally focused on the Disease-Free Equilibrium and the Reproductive Number. Other topics related to the mathematics of VSIR-like such as the honeymoon period (studied in [19]) have not been analyzed and could be covered in further research.

Basic properties of the model were stated, included the computation of the Reproduction Number and local asymptotically stability of the DFE. Then, a collection of propositions related to the optimization of the Reproduction number (including the Multiple Vaccination Theorem) were stated and proved. Two special results seem to be counterintuitive. Proposition 6 establishes that the limit when *r* → ∞ (the vaccination rate) is not zero if the leakiness is not zero. In theory, this limit value can be greater than one, and as a consequence, the vaccination campaign can not curtail the epidemic. Luckily, this situation was not observed in the selected countries.

Another important question is whether the addition of one type of vaccine helps in reducing the reproductive number. The answer to that question is that this situation depends on a key parameter *η* that acts as a threshold for the leakiness. Parameter estimation indicates that, for at least two countries, if this result was known and considered by the decision-makers, the reproductive number of the respective countries would have decreased even more. In addition, these vaccines could have been sent to other countries to reduce their reproduction number, which means that a better spatial distribution of the vaccines can be achieved in light of the new findings.

Finally, we should note that there exist some limitations for the application of this model in the data of COVID-19. As mentioned in the *Introduction*, this study is preliminary, and the authors aim to continue improving the model towards the consideration of more details that are particular for this pandemic.

## Data Availability

Data was collected from two different sources: Records from reported recovered and cumulative infected were taken from the John Hopkins repository [link 1] and for vaccination data, we used the repository of Our World in Data [link 2], including the datasets of vaccines per country and vaccines by the manufacturer.

https://github.com/CSSEGISandData/COVID-19

https://github.com/owid/covid-19-data/tree/master/public/data/vaccinations

## Declarations

### Conflict of Interest Statement

The authors have declared no competing interest.

### Funding

Ugo Avila Ponce de León is a doctoral student from Programa de Doctorado en Ciencias Biológicas of the Universidad Nacional Autónoma de México (UNAM). This paper was devoloped in the period of his PhD studies. Avila Ponce de León also received a fellowship (CVU: 774988) from Consejo Nacional de Ciencia y Tecnología (CONACYT). In addition, this work was supported in part by Universidad Autónoma de Yucatán and Mexican CONACYT under SNI grant numbers 15284.

### Availability of data and materials

Data was collected from two different sources: Records from reported recovered and cumulative infected were taken from the John Hopkins repository [35] and for vaccination data, we used the repository of *Our World in Data* [17], including the datasets of vaccines per country and vaccines by the manufacturer.

## Appendix

In this section, we provide additional and complementary results to the main propositions. Lemma 1 is the only one statement that is explicitly used in the text.

### Lemma 1.

*Let A, a matrix with all entries equal to zero but the values of the diagonal, the last row and the last column:*

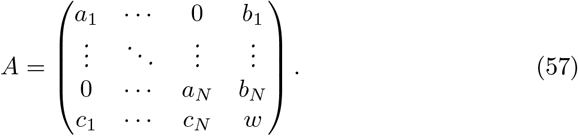

*Then*

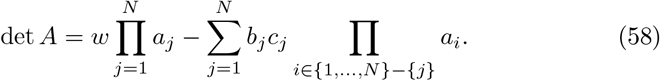

*Proof*. By Mathematical Induction on *N*, let consider the base case *N* = 1 with the matrix

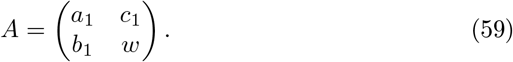

Then, det *A* = *wa*_1_ − *b*_1_*c*_1_, which satisfies (58). Now, let us consider the general case. Using twice the induction hypothesis,

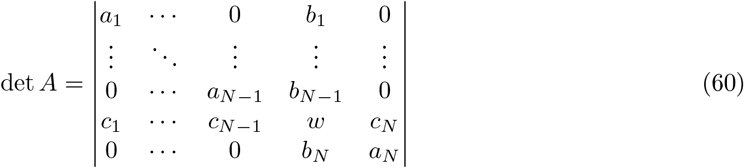

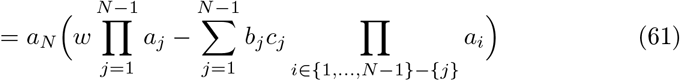

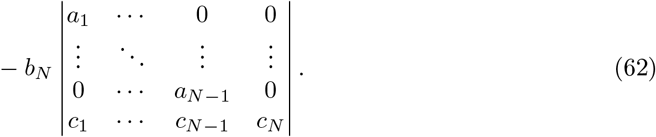

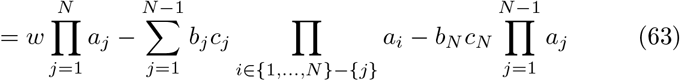

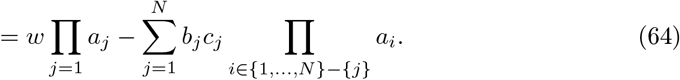

### Proposition 6.

*Let* ℛ_*N*_ *the Basic Reproduction Number of the model (1)-(4) with N vaccines and* ℛ_*N*+1_ *the Basic Reproduction Number of the model with two vaccines. Therefore, if ε*_*j,L*_ = 0 *for all j* = 1, …, *N* + 1, *then*, ℛ_*N*+1_ *<* ℛ_*N*_.

*Proof*. If *ε*_*j,L*_ = 0 holds, thus,

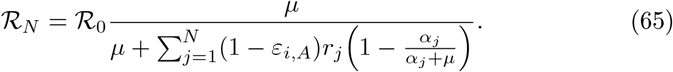

Since 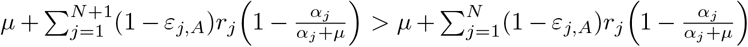, then,

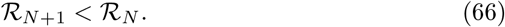

If the leakiness is set to 0, any new vaccine can help in reducing the reproduction number. Since the value of *η >* 0, this proposition is a particular case of the Multiple Vaccination Theorem.

### Proposition 7.

*Let ℛ*_1_ *the Basic Reproduction Number of the model (*1*)-(*4*) with one vaccine and ℛ*_*N*_ *the Basic Reproduction Number of the same model with N vaccines. let r*^1^ *the vaccination rate of the first case, and r*^2^ *the vaccination rate for all the j vaccines, and let p*^1^ *and p*^2^ *similarly defined for the parameter p*_*j*_. *If* 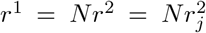, *and all vaccine parameters are equal (equivalents vaccines), then* ℛ_1_ = ℛ_*N*_.

### Proof.

Let 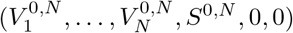 the DFE of the system with *N* vaccines and 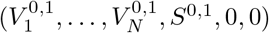 the DFE of the system with one vaccine. Using the hypothesis of the proposition, we get for all *j, ε*_*A*_ = *ε*_*j,A*_, *ε*_*L*_ = *ε*_*j,L*_, *α* = *α*_*j*_, and therefore,

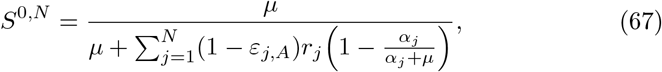

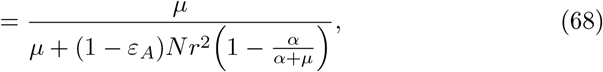

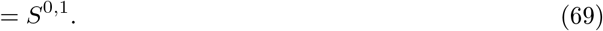

On the other hand,

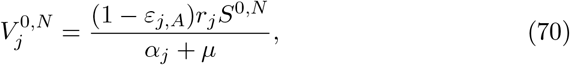

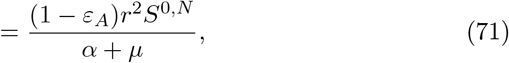

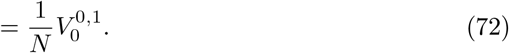

Then,

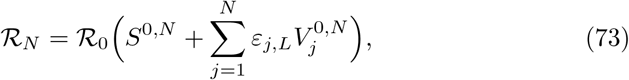

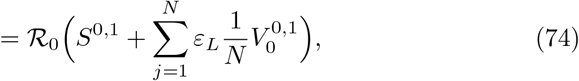

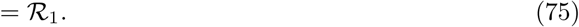

A direct consequence of Proposition 7, includes the case where *N* vaccines are not better than one: let consider a model with a single type with *r* as the vaccination rate. The same model, with *ε*_*L*_ = *ε*_*j,L*_ for all *j ≠ i* and *ε*_*L*_ *< ε*_*i,L*_, and the other vaccine parameters equal, *p* = *p*_*j*_ = 0 and *r* = *Nr*_*j*_ for all 0 *< j*≤ *N*. By Propositions 3 and 7, the model with *N* vaccines has a lower ℛ_*c*_. However, this case is not strange, because equivalent vaccines can be understood as the same one. More complex situations are covered in the main text, most of them explained by the Multiple Vaccination Theorem.

## Notes

### Author Declarations

IRB approval was not required since the work used only publicly available data.

